# Macrophage polarization in cesarean scar diverticulum

**DOI:** 10.1101/2021.11.25.21266862

**Authors:** Jinfa Huang, Xiaochun Liu, Yi Hou, Yixuan Liu, Kedan Liao, Ning Xie, Kaixian Deng

## Abstract

**Aims:** To determine immunohistochemical features and correlations between M1/M2 polarization status with disease severity of post-cesarean scar diverticulum (CSD).

**Methods:** Histological and immunohistological staining were performed and inflammatory (CD16, CD163, and TNF-α), fibrosis (α-SMA), and angiogenic (CD31) markers were examined in uterine tissues collected from patients with uterine scar diverticula (CSD) (n=37) and cesarean section (CS) (n=3).

**Results:** CSD tissues have higher expression of α-SMA, TNF-α, CD16, and CD31 and lower expression of CD163 than CS tissue (P <0.05). Compared with adjacent tissues, thick-walled blood vessels, glands, and fibrotic sites have higher expression of α-SMA, TNF-α, and CD16. Statistical correlation was observed between the expression of CD16 and TNF-α (R = 0.693, P <0.001), α-SMA (R = 0.404, P <0.05), and CD31 (R = 0.253, P <0.05) in CSD tissues, especially with the ratio of CD16/CD163 (R = 0.590, P <0.01). A more significant difference was observed between the expression of CD16/CD163 and α-SMA (R = 0.556, P <0.001), TNF-α (R = 0.633, P <0.0001) and CD31 (R = 0.336, P <0.05) Statistical correlation.

**Conclusion:** In this study, TNF-α, α-SMA, CD16, and CD31 proteins were overexpressed in all CSD cases, and CD16/CD163 was positively correlated with tissue inflammation, fibrosis, and neovascularization. Abnormal mononuclear macrophage infiltration may be involved in the origin and progression of CSD.

## Introduction

Cesarean scar diverticulum(CSD)also known as uterine niches, uterine pouches, or diverticula, is characterized by a recess in the lower uterine segment due to healing defects after cesarean section[1]. According to reports, the incidence of CSD varies from 24% to 84% through transvaginal ultrasound or hysteroscopy[2]. Among them, 20–25% are asymptomatic, while others may develop common symptoms such as abnormal uterine bleeding, excessive menstruation, prolonged menstrual or bleeding, dysmenorrhea, chronic pelvic pain, and infertility[3-6]. The pathological mechanism and pathological characteristics that lead to these clinical symptoms of CSD have not been reported.

Macrophages are vital for regulating inflammation, tissue repair, regeneration, and fibrosis[7]. In the local tissue microenvironment, macrophages of different phenotypes exhibit distinct heterogeneity in response to stimuli and cytokines release[8]. The two classic macrophages include typically activated macrophages (M1 macrophages) induced by LPS and IFN-γ, and Th2 cytokines, such as IL-4 and IL-13 induced alternate activation of macrophages (M2 macrophages)[9]. Other M2 phenotypes can be induced by MCSF and IL-10[10]. Uterine wound repair is generally characterized as an inflammatory and wound-healing response, where these two phenotypes of macrophages are dominant[11]. After tissue injury, a large number of macrophage precursors are recruited to the injury site, which not only acts as scavenger cell but also produces chemokines and inflammatory cytokines to recruit more inflammatory cells[12]. In the fading phase of inflammation, M2 phenotypic macrophages dominate and promote cell proliferation and vascular development by up-regulating many anti-inflammatory molecules and growth factors[13]. When the balance of M1/M2 macrophages is disrupted, the tissue microenvironment is disturbed, resulting in abnormal tissue repair, regeneration, and fibrosis. Studies have shown that macrophages are involved in the damage, repair, regeneration, and fibrosis of the lung, liver[14], kidney[15], skin, and other tissues[16].

The formation of CSD is more complicated than uterine wound healing. However, there are limited relevant reports, so it seems obvious that more basic knowledge is needed for a better understanding of the development and progress of the disease. In this study, we aim to characterize the morphology and pathology of CSD, involving histological changes and macrophages infiltration, as well as its association with fibrosis, inflammation, and angiogenesis.

## Material and methods

We prospectively recruited patients with uterine scar diverticulum who underwent surgical treatment for reproductive needs. Samples from women who underwent total hysterectomy for uterine fibroids (with a history of cesarean section surgery) serve as a reference. In all patients, at least 1×1cm tissues should be collected from the surgically resected specimen and fixed in formalin solution for subsequent experiments. The study was approved by the ethics committee of Shunde Hospital of Southern Medical University, and everyone signed a written informed consent before surgery.

Haematoxylin-eosin (HE) staining and immunohistochemical staining Immunohistochemistry staining and haematoxylin-eosin (HE) staining were performed as described in the previous research[17]. The paraffin-embedded tissue was cut into 4 μm-thick sections and then stained for histological evaluation and immunohistochemical analysis. The primary antibodies are as follows: 1:200 diluted polyclonal mouse anti-human CD31 antibody, α-SMA antibody, CD16 antibody, CD163 antibody, TNF-α antibody (Sevier; China)[18].

### Histological evaluation

H-score: the abbreviation of histochemistry score, is a comprehensive histological score of the number of positives in each section and the intensity of staining, to achieve the goal of semi-quantitative tissue staining. H-SCORE=Σ(pi×i)=(percentage of weak intensity area ×1)+(percentage of moderate-intensity area ×2)+(percentage of strong intensity area ×3), where pi means positive Signal pixel area percentage; i represents the positive level. The H-score is a value between 0-300. The larger the value, the stronger the overall positive intensity.

### Statistical analysis

The statistical software package IBM SPSS Statistics version 20.0 (IBM Inc., Chicago, IL, USA) is used for statistical analysis. Paired t test and chi-square test were used to compare differences between groups. P value < 0.05 was considered significant.

## Results

### General clinical information

There was no significant difference in age between patients with CSD (mean: 28.0 years, range: 21-42) and those with CS (mean: 29.3 years, range: 26-35) (P > 0.05). 100% of the lesion is located in the lower part of the uterus (Table S1).

### Histological characteristics

The histological characteristics of H&E staining are summarized in Table S2 and depicted in Figure S1. In 100% (37/37) CSD patients, endometrial and muscular layer defects were observed, but no obvious muscular layer defects and 33.33% (1/3) endometrial defects were seen in CS patients. Most patients have reduced or atrophied glands (81.08% of patients with diverticula and 66.67% of patients with CS), but dilated gland is only seen in patients with CSD (45.95%). The proportion of fibrosis (89.19% vs 33.33%) and thick-walled blood vessels (78.38 vs 33.33%) in patients with CSD is higher than that of patients with CS. In addition, endometrial atypia (54.05%) and congested capillaries (86.49%) around the diverticulum are only present in patients with CSD (Figure S1).

### Immunohistochemical characteristics

#### Fibrosis analysis of uterine scar diverticulum

The fibrosis extent of the uterine tissue was assessed by α-SMA staining. All patients had α-SMA immunoreactivity. The H score of CSD tissue was higher than that of the control group (p < 0.05). The α-SMA is mainly located in the cytoplasm. The proportion of a positively stained area of α-SMA in CSD tissue is 33%-81%. And the intensity of α-SMA staining at the proximal end of the diverticulum tissue was lower than that of the distal part. In addition, around thick-walled blood vessels, glands, and fibrotic tissues, α-SMA staining is strongly positive (Figure 1). Analysis of levels of inflammation and inflammatory cell infiltration of CSD

**Figure 1.**
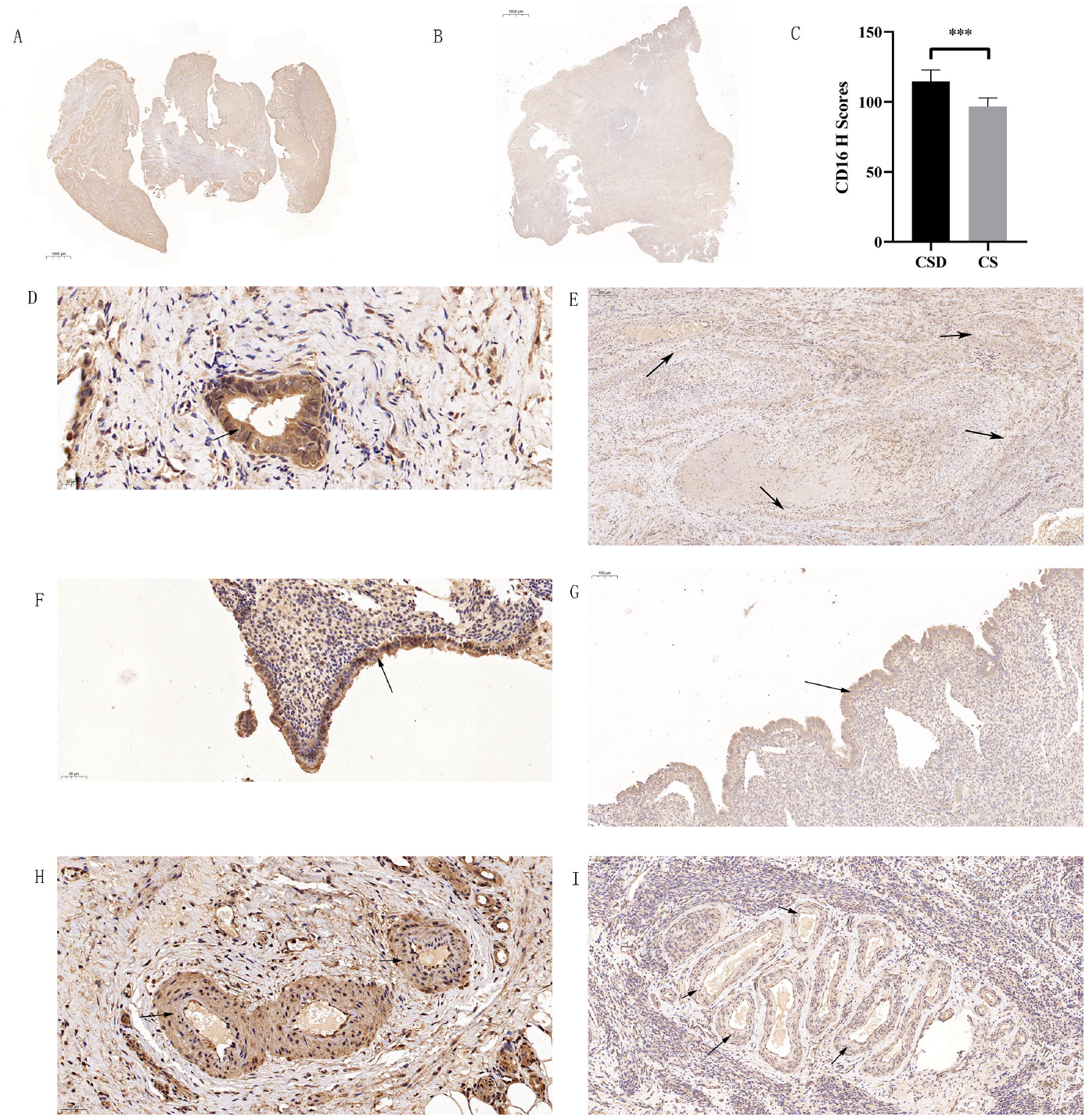
Representative immunohistochemical characteristics of α-SMA in CSD and CS tissues. (A) A representative image of α-SMA in CSD. (B) A representative image of α-SMA in CS tissue. (C) The quantitative analysis of α-SMA immunoreactivity using H-score. * p < 0.05. (D) A representative image of α-SMA in the thick-walled blood vessel in CSD. (E) α-SMA in the thick-walled blood vessel in CS. (F) α-SMA in the gland in CSD. (G) α-SMA in the gland in CS. (H) α-SMA in fibrotic tissues in CSD. (I) α-SMA in fibrotic tissues in CS. The scale bar is shown in the picture. The arrow indicates the positive staining area.

#### Analysis of inflammation of CSD

The inflammation levels of the uterine tissue were assessed by TNF-α staining. All patients with CSD and the control group had different degrees of immunoreactivity to TNF-α (Figure 2). The H score of CSD tissue was higher than that of the control group (p < 0.05). TNF-α staining is located in the cytoplasm and nucleus. The positive area ratio of TNF-α in CSD tissue ranges from 30% to 82%. And the intensity of TNF-α staining at the proximal end of the diverticulum tissue was higher than that of the distal areas. In addition, around thick-walled blood vessels, TNF-α staining is strongly positive.

**Figure 2.**
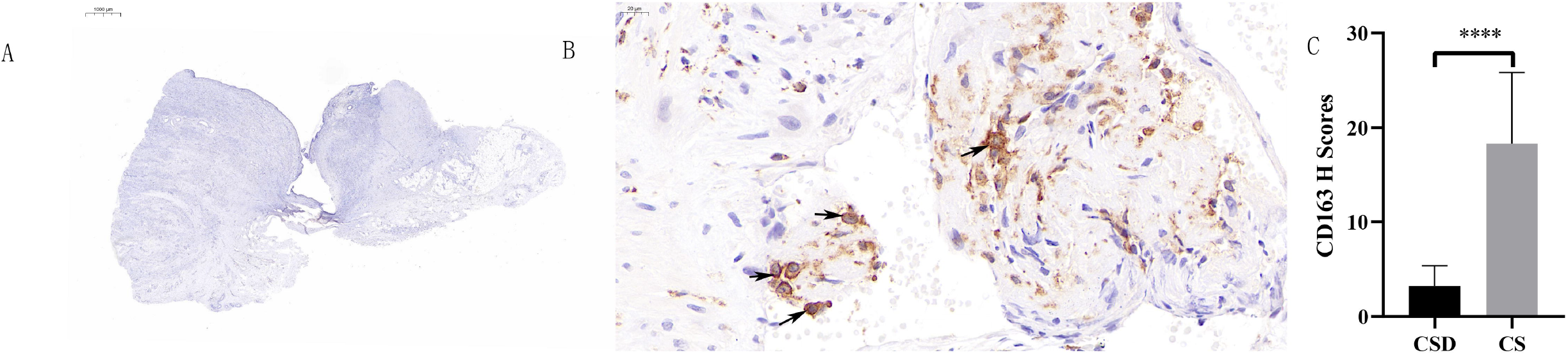
Representative immunohistochemical characteristics of TNF-α in CSD and CS tissues. (A) A representative image of TNF-α in CSD. (B) A representative image of TNF-α in CS tissue. (C) The quantitative analysis of TNF-α immunoreactivity using H-score. * p < 0.05. (D) A representative image of TNF-α in the thick-walled blood vessel in CSD. (E) TNF-α in the thick-walled blood vessel in CS. (F) TNF-α in the glands in CSD. (G) TNF-α in the glands in CS. (H) TNF-α in the endometrium in CSD. (I) TNF-α in the endometrium in CS. (J) TNF-α of capillaries in CSD. (K) TNF-α of capillaries in CS. The scale bar is shown in the picture. The arrow indicates the positive staining area.

#### Inflammatory cell infiltration of CSD

The inflammation cell infiltration of the uterine tissue was assessed by CD16 (M1)[19] and CD163 (M2)[20] staining. All CSD have CD16 immunoreactivity, while CD16 in the control group is weakly positive or negative. The H score of CSD tissue was higher than that of the control group (p < 0.05). CD16 is located in the cytoplasm and nucleus. The CD16 positive area ratio of CSD tissue ranges from 31% to 59%. And the intensity of CD16 staining at the proximal end of the diverticulum tissue was higher than that at the distal end. In addition, CD16 staining is strongly positive around thick-walled blood vessels (Figure 3).

**Figure 3.**
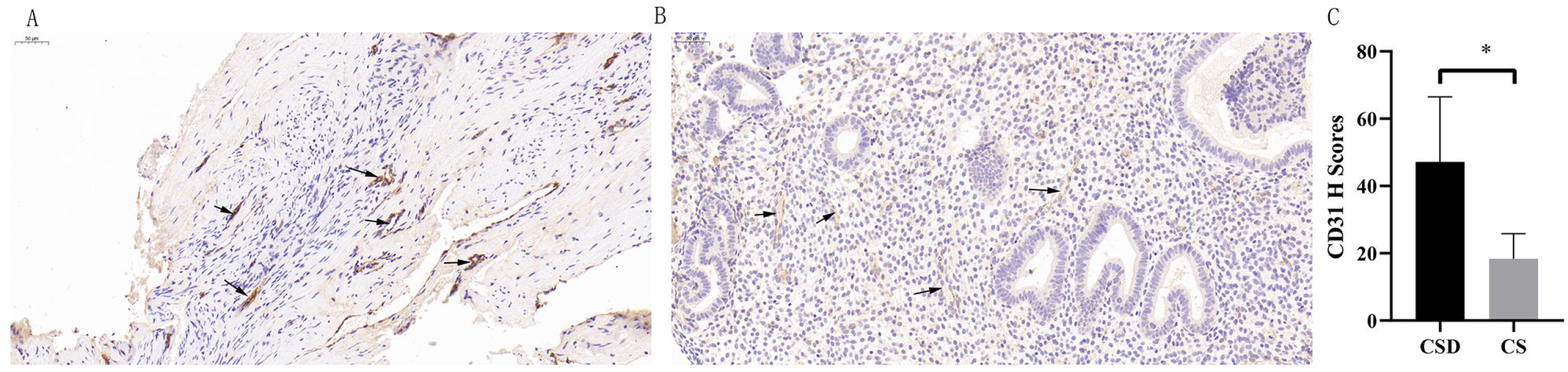
Representative immunohistochemical characteristics of CD16 in CSD and CS tissues. (A) A representative image of CD16 in CSD. (B) A representative image of CD16 in CS. (C) The quantitative analysis of CD16 immunoreactivity using H-score. *** p < 0.001. (D) A representative image of CD16 in the glands in CSD. (E) CD16 in the glands in CS. (F) CD16 in the endometrium in CSD. (G) CD16 in the glands in CS. (H) CD16 in the thick-walled blood vessel in CSD. (I) CD16 in thick-walled blood vessel in CS. The scale bar is shown in the picture. The arrow indicates the positive staining area.

In addition, the H scores of CD163 in CS were higher than that of CSD (P < 0.05) (Figure 4), and the H scores of CD31 in CSD were higher than that of the control group(P < 0.05) (Figure 5).

**Figure 4.**
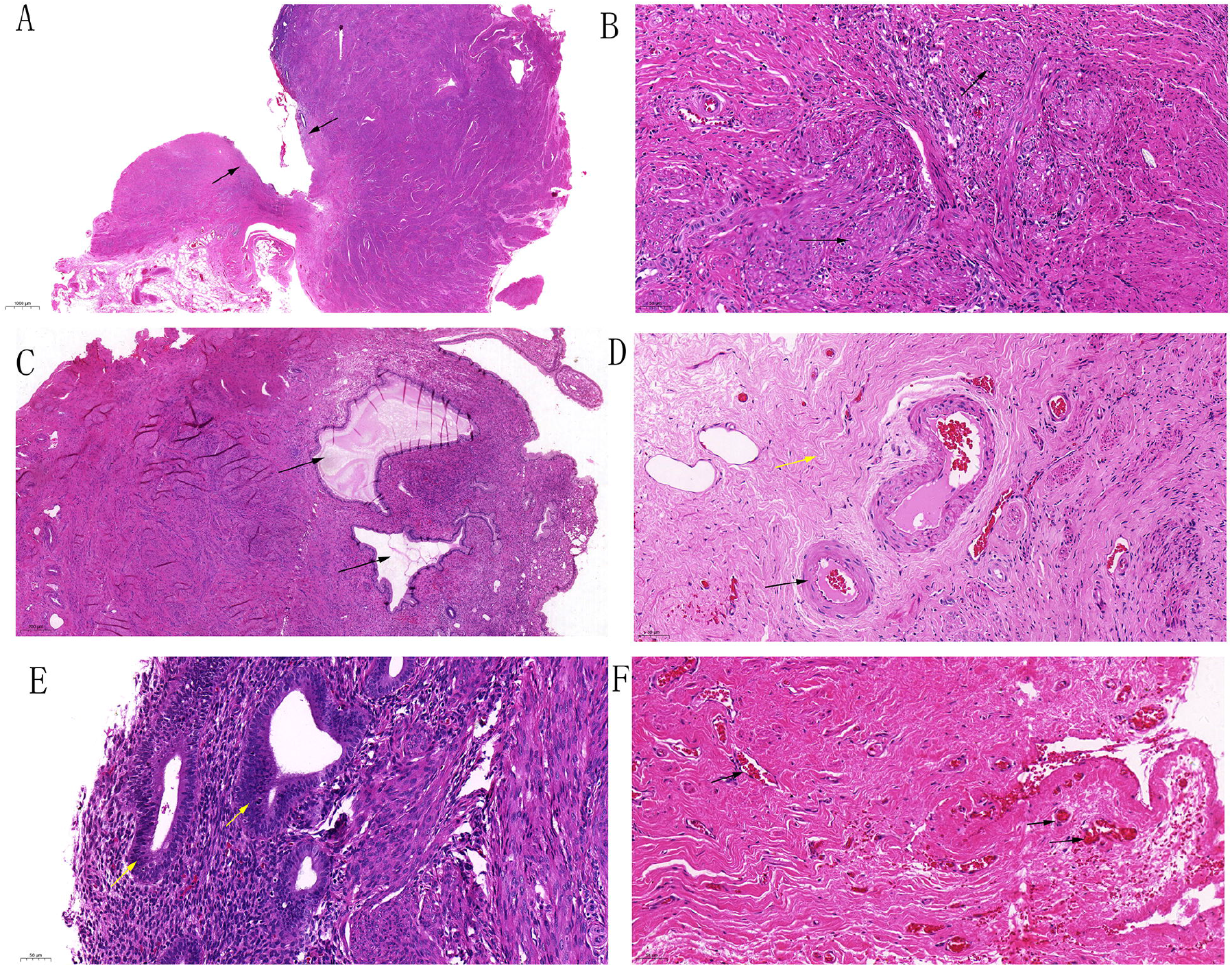
Representative immunohistochemical characteristics of CD163 in CSD and CS tissues. (A) A representative image of CD163 in CSD. (B) Representative image of CD163 in CS tissue. (C) The quantitative analysis of CD163 immunoreactivity using H-score. **** p < 0.0001. The scale bar is shown in the picture. The arrow indicates the positive staining area.

**Figure 5.**
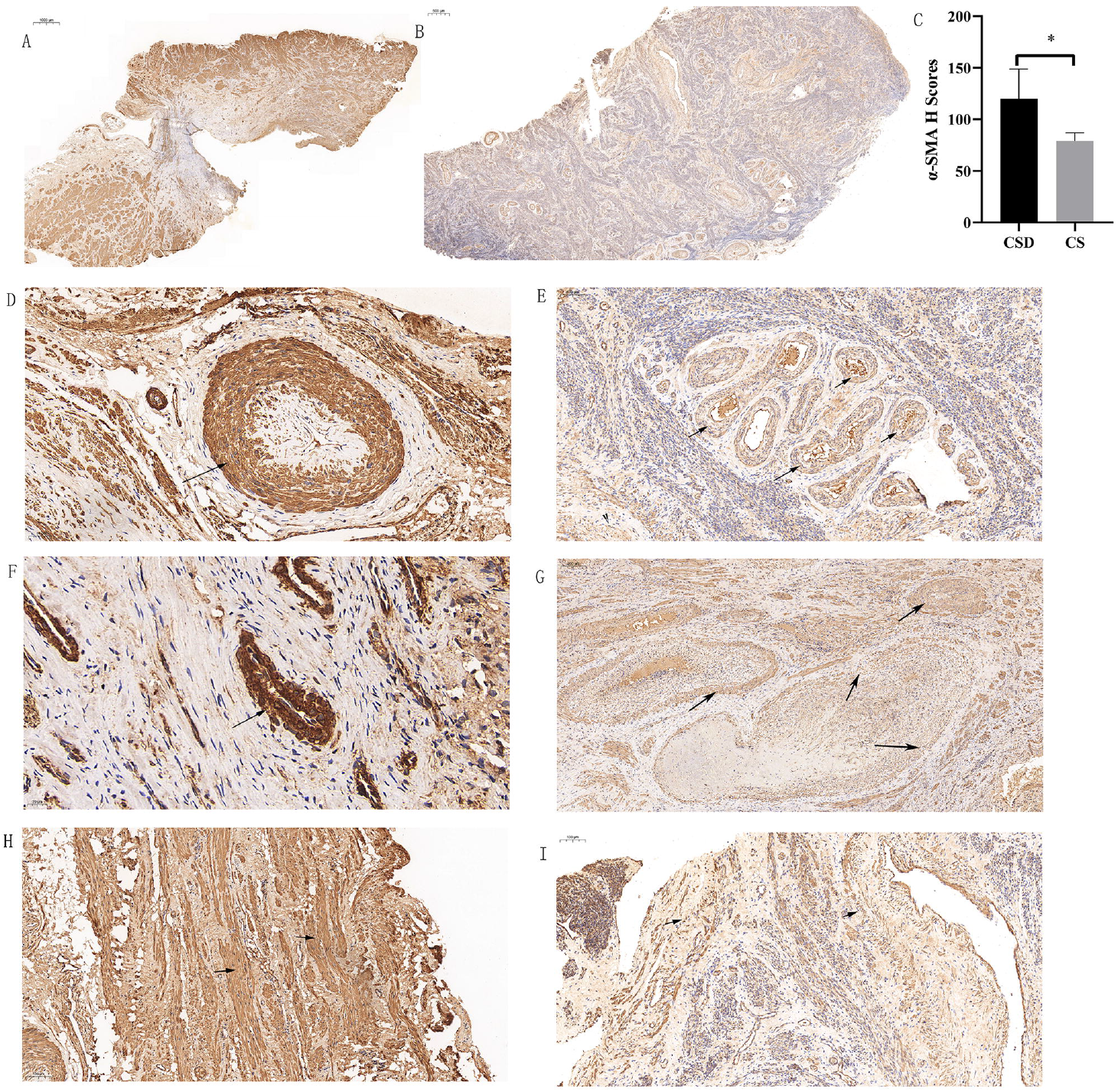
Representative immunohistochemical characteristics of CD31 in CSD and CS tissue. (A) A representative image of CD31 in CSD. (B) A representative image of CD163 in CS. (C) The quantitative analysis of CD31 immunoreactivity using H-score. * p < 0.05. The scale bar is shown in the picture. The arrow indicates the positive staining area.

Correlation analysis between α-SMA, TNF-α, CD31, CD16, CD163, and CD16/CD163 expression in CSD tissues of immunohistochemical staining

We assessed the correlation between α-SMA, TNF-α, CD31, CD16, CD163, and CD16/CD163 by Spearman correlation analysis in uterine tissues of 37 CSD patients (Table 1). Statistically positive correlations were observed between α-SMA and TNF-α expression (R = 0.500, P < 0.01), between α-SMA and CD16 expression (R = 0.404, P < 0.05), between α-SMA and CD16/CD163 expression (R = 0.556, P < 0.001), between TNF-α and CD16 expression (R = 0.693, P < 0.001), between TNF-α and CD31 expression (R = 0.272, P < 0.05), between TNF-α and CD16/CD163 expression (R = 0.633, P < 0.0001), between CD16 and CD31 expression (R = 0.253, P < 0.05), between CD16 and CD16/CD163 expression (R = 0.590, P < 0.01), between CD31 and CD16/CD163 expression (R = 0.336, P < 0.05) in CSD. No statistical correlations were observed between CD31 and α-SMA expression, between CD163 and TNF-α, CD16, as well as CD31 expression in CSD. Interestingly, negative correlations between the expressions of α-SMA and CD163 (R = -0.428, P < 0.05), CD163 and CD16/CD163 (R = -0.672, P < 0.01) were observed in 37 CSD tissues.

**Table 1.** Correlation among α-SMA, TNF-α, CD16, CD31, CD163 and CD16/CD163 expression in CSD tissues.

| | α-SMA | TNF-α | CD16 | CD31 | CD163 |
| --- | --- | --- | --- | --- | --- |
| TNF-α | 0.500** | | | | |
| CD16 | 0.404* | 0.693*** |  |  |  |
| CD31 | ns | 0.272* | 0.253* |  |  |
| CD163 | -0.428* | ns | ns | ns |  |
| CD16/<br>CD163 | 0.556*** | 0.633**** | 0.590** | 0.336* | -0.672** |

## Discussion

This study emphasized the immunohistochemical characteristics of CSD. To our knowledge, there have been no reports of immunohistochemical studies on CSD. A large number of mononuclear cell infiltration (immunoreactive to CD16), higher levels of tissue inflammation (immunoreactive to TNF-α), and fibrosis levels (immunoreactive to α-SMA) were observed in the tissues of CSD, as well as more new blood vessels (immunoreactive to CD31) and almost non-existent M2 (immunoreactive to CD163) macrophages. In the tissue distribution, the degree of fibrosis in the distal uterine scar diverticulum lesions is higher than that in the proximal tissue, and the proximal tissue has a higher level of mononuclear cell infiltration and tissue inflammation.

In addition, this study also emphasized the histomorphological characteristics of CSD. We observed endometrial atypia, expanded glands, and hyperemia of capillaries around the CSD, which are the unique characteristics of CSD. In addition, fibrosis and thick-walled blood vessels are more common. These specific tissue structures present unique immunohistochemical staining characteristics, such as increased fibrosis around thick-walled blood vessels, stronger CD31 staining of hyperemic capillaries, and more CD16-positive cells around the atypical endometrium. These tissue morphological characteristics have been reported in previous studies[21].

Macrophages and the cytokines secreted by them are key regulators and participants in wound healing. The imbalance of immune response during wound healing, such as macrophage polarization disorder, will cause the inflammation phase to fail to subside, which will lead to poor and delayed wound healing[22, 23]. Specifically, there are too many pro-inflammatory macrophages, while the number of macrophages with anti-inflammatory and tissue damage repair phenotypes is limited[24, 25]. In CSD, we observed a large number of monocyte infiltration and limited M2 cell infiltration, suggesting that macrophage polarization disorder (M1 phenotype conversion to M2 phenotype) is closely related to the pathology of the disease. In addition, the reduced ability of macrophages to clear dead neutrophils is another characteristic of chronic wounds. This leads to the persistence of tissue microinflammation, and these cells produce a large number of inflammatory mediators, such as tumor necrosis factor-α (TNF-α) and interleukin-1β (IL-1β)[25]. Consistent with chronic wounds, there is a large amount of TNF-α in the CSD. In addition to the accumulation of pro-inflammatory macrophages, the possible cause may also be impaired clearance of apoptotic neutrophils by macrophages. Inflammatory cell infiltration and a large number of inflammatory factors lead to delayed or non-healing wound healing, which can cause long-term retention of menstrual blood in the wound defect site, and even cause infection, which will further increase the level of tissue inflammation and hinder the progress of wound healing.

Excessive fibrosis is another characteristic of CSD. Emerging evidence suggests that macrophages are inflammatory (M1) and the alternatively activated macrophages (M2) participate in the pathogenesis of various fibrotic diseases[26, 27]. M1 macrophages are involved in fibrosis. In contrast, M2 macrophages secrete IL-10 and arginase-1 and exert their anti-inflammatory properties[28, 29]. Therefore, the polarization of tissue macrophages is the key to regulating tissue fibrosis. In this study, we found that macrophage polarization disorder is closely related to the degree of tissue fibrosis.

A large number of new capillaries is a common feature of many chronic inflammatory diseases. There are a large number of new blood vessels in CSD, which suggests that it also has the characteristics of chronic wound healing. It is reported that macrophages promote angiogenesis by secreting pro-angiogenic factors, such as tumor necrosis factor-α (TNF-α) and endothelial growth factor (VEGF)[32]. Inflammatory cell infiltration and TNF-α in the tissue may be the reason for the formation of many new blood vessels in the CSD. In addition, pro-inflammatory cytokines, chemokines, and other inflammatory mediators released by inflammatory cells can also cause new blood vessels.

## Conclusion

This study is helpful to understand the pathology and molecular mechanism of CSD. CSD has a wide range of macrophage polarization abnormalities, persistent tissue micro-inflammation, fibrosis, and increased histological characteristics of new blood vessels, which induce characteristic tissue structures including endometrial atypia. Abnormal macrophage polarization states are positively correlated with tissue inflammation level, fibrosis, and angiogenesis in patients with CSD. Therefore, it is necessary to further study the causes of abnormal polarization of macrophages in CSD and use them as prognostic factors and therapeutic targets.

## Data Availability

The authors declare that the data and material of this study are available within the article.

## Authors’ contributions

Kaixian Deng conceptualized the study design and supervised the analysis. Jinfa Huang conducted the experiments, performed the statistical analysis, and wrote the paper; Xiaochun Liu prepared the samples; Yi Hou revised the manuscript; Yixuan Liu, Ning Xie and Kedan Liao provided supervision of the analysis and prepared the article. All authors read and approved the final manuscript.

## Acknowledgements

Not applicable.

## Compliance with ethical standards

The authors declare that they have no competing interests.

## Availability of data and materials

The authors declare that the data and material of this study are available within the article.

## Take home messages

Immunohistochemical features and macrophage infiltration levels was analysed in post-cesarean scar diverticulum(CSD).

Dysregulated macrophage polarization and high levels of tissue inflammation, fibrosis and abnormal angiogenesis were observed in CSD. Macrophage polarization correlates with tissue inflammation, fibrosis and abnormal angiogenesis and may be linked with disease progression in CSD.

---

**Figure S1.**
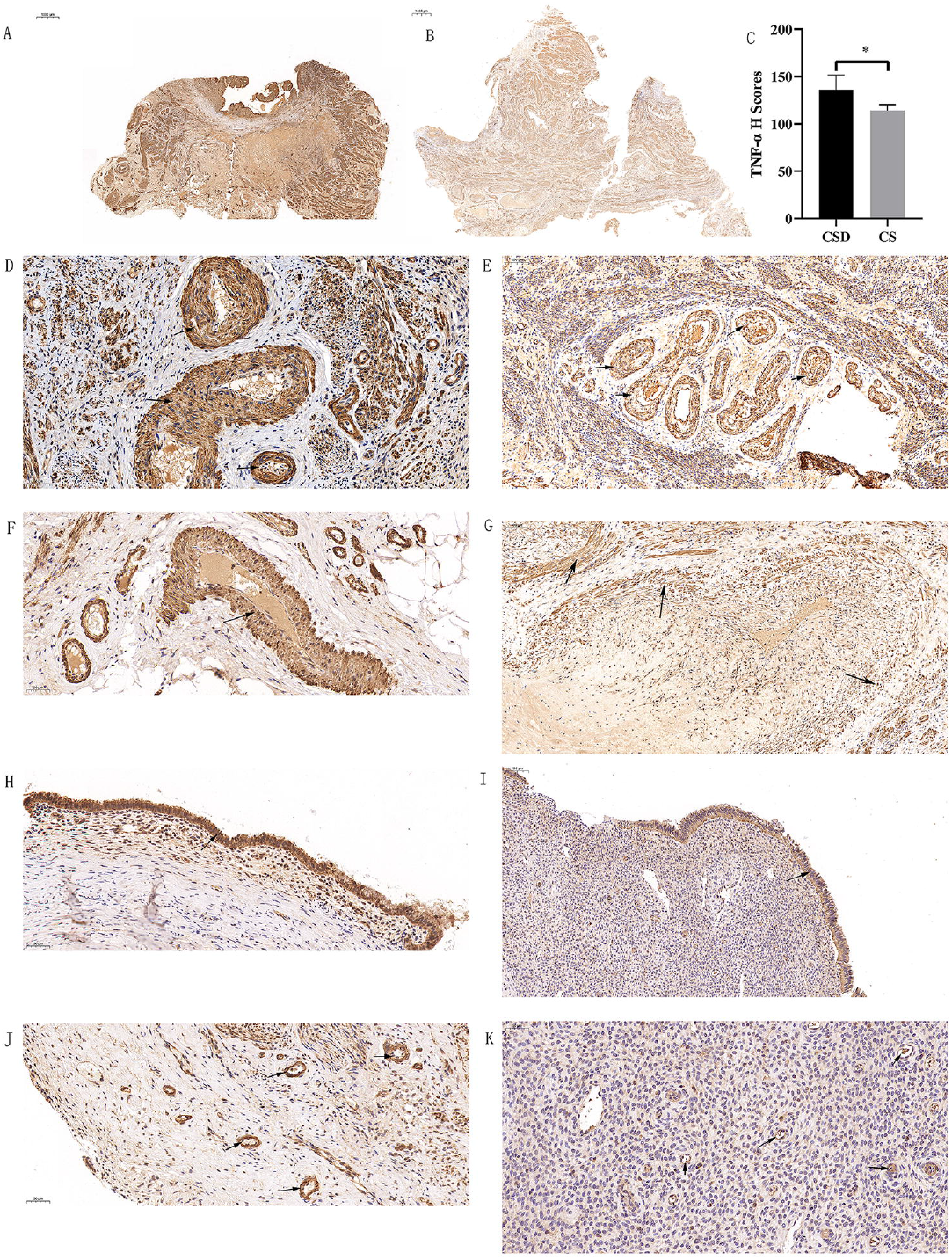
Representative histomorphological characteristics of CSD and CS tissues. (A) Pictures of uterine scar diverticulum. (B) Atrophic gland tissue. (C) Dilated glands. (D) Fibrosis and thick-walled blood vessels. (E) Endometrial atypia. (F) Hyperemia of capillaries. The scale bar is displayed in the bottom left of the picture. The arrow indicates the characteristic structures.

**Table S1.** Clinic characteristics of CSD and CS patients.

|  | CSD | CS | P value |
| --- | --- | --- | --- |
| Age | 28.3±3.27 | 29.3±4.93 | P>0.05 |
| Cesarean section location<br>(lower part of the uterus) | 100% | 100% | P>0.05 |

**Table S2.** The morphological characteristics of CSD and CS tissues.

|  | CSD (n=37) | CS (n=3) |
| --- | --- | --- |
| Endometrial defects | 100%(37) | 33.33%(3) |
| Muscular layer defects | 100%(37) | 0 |
| Reduced or atrophied glands | 81.08%(30) | 66.67%(2) |
| Dilated gland | 45.95%(17) | 0 |
| Fibrosis | 89.19%(33) | 33.33%(1) |
| Thick-walled blood vessels | 78.38%(29) | 33.33%(1) |
| Endometrial atypia | 54.05%(20) | 0 |
| Congested capillaries | 86.49(32) | 0 |

